# Determining the optimal hematoma volume-based thresholds for surgical and medical strategies in basal ganglia hemorrhage

**DOI:** 10.1101/2024.06.23.24309367

**Authors:** Chonnawee Chaisawasthomrong, Atthaporn Boongird

**Affiliations:** Division of Neurosurgery, Department of Surgery, Ratchaburi Hospital, Ratchaburi, Thailand; Division of Neurosurgery, Department of Surgery, Faculty of Medicine Ramathibodi Hospital, Mahidol University, Bangkok, Thailand

**Author notes:** **Corresponding author:** Assoc.Prof. Atthaporn Boongird MD, Head section of division, Division of Neurosurgery, Department of Surgery, Ramathibodi Hospital, Mahidol University, Bangkok 10400,Thailand #256.

**Keywords:** Basal ganglia hemorrhage, hematoma volume, medical treatment, surgical intervention

## Abstract

**Background:** The indication for surgical intervention in spontaneous intracerebral hemorrhage remains controversial, particularly regarding the benefits of early hematoma drainage via open craniotomy. This study aimed to identify the maximum hematoma volume suitable for conservative treatment and the volume that represents an absolute indication for surgery in patients with basal ganglia hemorrhage.

**Methods:** A retrospective analysis was performed on the medical records of patients admitted for basal ganglia hemorrhage from 2019 to 2021. The data encompassed personal history, general information and diagnostic imaging records, particularly CT brain scans from the initial ER visit, were examined to ascertain hematoma volume. The comparison focused on evaluating the outcomes of patients who received medical treatment compared to those who underwent surgical intervention, mainly considering various hematoma volumes, and was conducted using multivariate logistic analysis.

**Results:** In a study of 387 cases of basal ganglia hemorrhage, analysis of medical treatment alone across various hematoma volumes revealed that the group with volumes between 10 and 39.9 ml showed no significant difference in mortality compared to the group with volumes less than 10 ml. The Receiver Operating Characteristics (ROC) curve identified a 45.3 ml cutoff for survival prediction with medical treatment alone. Notably, patients in the subgroup undergoing surgical intervention with a hematoma volume less than 30 ml exhibited significantly higher mortality than those who did not undergo surgery. Conversely, there was a pronounced and statistically significant trend toward increased survival in the group with a hematoma volume of at least 60 ml.

**Conclusions:** The application of medical treatment alone is suitable for hematoma volumes ranging from 0 to 45.3 ml, whereas volumes of 60 ml or more serve as a clear indication for surgical intervention in patients with basal ganglia hemorrhage.

## Introduction

Spontaneous intracerebral hemorrhage (SICH) is a common occurrence, with an annual incidence ranging between 11 and 23 cases per 100,000^(1)^. In the United States, there were 107,590 identified discharges related to ICH, and a craniotomy procedure code was detected in 7,518 instances (7.0%)^(2)^. The mortality rate for ICH within 30 days is 40% to 50%, which is double that of ischemic stroke. Only 27% of patients achieve functional independence after 90 days ^(3),(4)^. Following the evacuation of intracranial hemorrhage, the 30-day mortality rate was 23.3% ^(5)^. Notably, mortality was higher among individuals aged 65 years and older (p = 0.020). The deep-seated group exhibited a higher incidence and extent of intraventricular extension, as well as a younger age (52.6 ± 9.0 years vs. 58.5 ± 9.8 years; p < 0.05)^(6)^. The indication for surgical intervention for SICH remains controversial^(10)^. Some studies suggest that surgery is warranted when the hematoma volume exceeds 30 ml^(7)^. In cases of secondary neurological deterioration with a hematoma volume ranging from 50 to 60 ml, performing an open craniotomy and evacuating the hematoma can be life-saving^(8)^. In clinical practice, there were numerous patients presenting to the emergency room with more than 30 ml of SICH, yet their neurological examination was intact, and they could survived without further surgical intervention. The role of open craniotomy for early hematoma drainage after intracranial hemorrhage remains a topic of hot debate, and randomized controlled trials have failed to demonstrate its benefit in terms of mortality or functional outcomes ^(9)^. The STICH II results confirm that early surgery does not increase the rate of death or disability at 6 months and might offer a small but clinically relevant survival advantage for patients with spontaneous superficial intracerebral hemorrhage without intraventricular hemorrhage ^(11,12)^.

This paper aimed to conduct a retrospective comparison between medical treatment and surgical intervention in basal ganglia hemorrhage across different volumes. The objective is to identify the maximum volume suitable for conservative treatment and the volume that serves as an absolute indication for surgery.

## Methods

A retrospective data collection was performed on patients diagnosed with basal ganglia hemorrhage at Ratchaburi Hospital in Ratchaburi Province, Thailand, between the years 2019 and 2021. The patients were closely monitored for a period of 30 days following the occurrence of intracerebral hemorrhage during their hospital stay. Any incomplete medical records, patients who demonstrate resistance towards seeking hospital care or exhibit attempts to evade medical treatment, instances of intracerebral hemorrhage attributed to secondary causes such as a bleeding tumor, ruptured aneurysms, arteriovenous malformation, arteriovenous fistula, hemorrhagic transformation, venous sinus thrombosis, coagulopathy and patients who experienced hematoma expansion during treatment, were excluded from the research analysis. History and diagnostic imaging results were gathered from all medical records. Personal history and general information, such as gender, age, underlying diseases, alcohol consumption, and smoking habits, were also recorded. Additionally, The Radiographic imaging history included volume of hematoma, interpreted by the radiologists using Synapse software version 4.4.210 (Fujifilm Thailand, Bangkok, Thailand).

The volume of the hematoma within the basal ganglia was calculated without including intraventricular hemorrhage volume, using the formula A × B × C / 2. Here, A represents the greatest hemorrhage diameter on the CT axial view, B represents the diameter perpendicular to A, and C represents the maximal diameter of the hematoma on the coronal view Figure 1). The data analysis was performed using Stata software version 17 (StataCorp, College Station, TX, USA).

**Figure.1.**
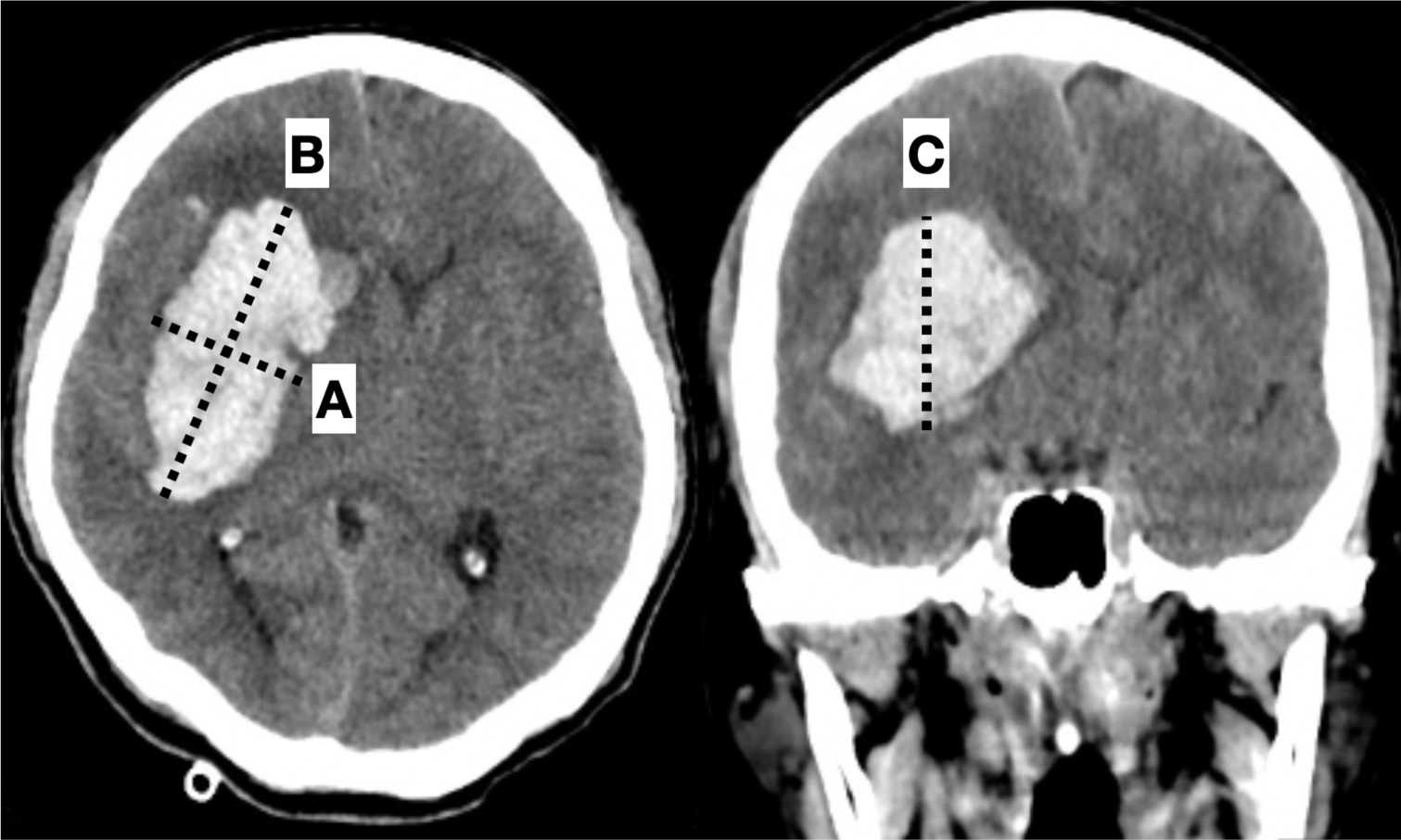
Volume of the hemorrhage A x B x C/2. A dash line, the greatest hemorrhage diameter on the CT axial view; B dash line, the diameter perpendicular to A dash line; C dash line, the maximal diameter of the hemorrhage on the coronal view.

An analysis was undertaken to determine the maximum volume that could be managed through medical treatment by comparing outcomes among patients who either survived or died while receiving medical treatment across various hematoma volumes. Simultaneously, the study aimed to identify the minimum volume that could be effectively treated with surgical intervention, minimizing morbidity and mortality. This comparison involved assessing the mortality outcomes of patients who received medical treatment against those who underwent surgical intervention at different hematoma volumes. The goal was to establish the threshold volume for medical treatment associated with survival, and the minimum volume necessitating surgical intervention while minimizing adverse outcomes, focusing solely on the volume of hematoma without considering clinical manifestations of consciousness or signs of herniation.

For variables with a normal distribution, Students t-test or Mann-Whitney U test were utilized. Univariate and multivariate logistic regression analysis were employed for confounding data. A significance level of P < 0.05 was considered statistically significant.

## Results

In Ratchaburi Hospital from 2019 to 2021, a total of 387 cases of basal ganglia hemorrhage were identified. Among these cases, 132 patients (34.11%) underwent surgical intervention, while 255 patients (65.89%) received only medical treatment. The mean age for patients undergoing surgical intervention was 56.86±12.26, and for those receiving medical treatment, it was 58.65±13.36. There was no significant difference in mean age between the two groups (OR 0.99; 0.97-1.00; p = 0.200). Regarding gender, 91 males (33.21%) underwent surgery compared to 41 females (36.28%). Additionally, 183 males (66.79%) received only medical treatment, while 72 females (64.72%) did so. However, these gender differences were not statistically significant (OR 0.87; 95%CI 0.55-1.38; p = 0.562). Factors including coronary heart disease (CAD), chronic kidney disease (CKD), smoking, and alcohol habits were considered potential confounding factors. There were no statistically significant differences in these parameters between the surgical intervention and medical treatment groups (Table 1).

**Table. 1.**
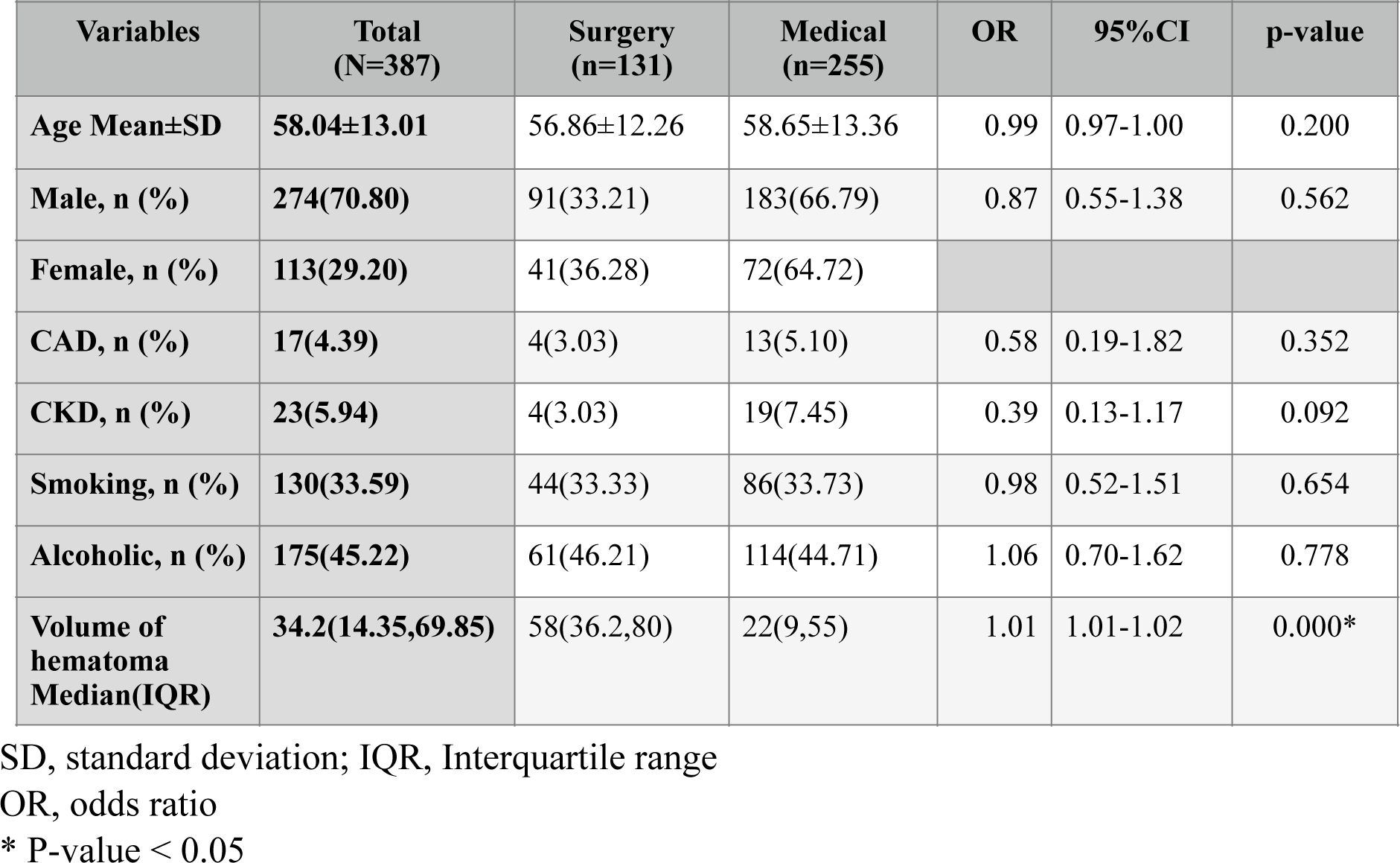
Demographic Data for Patients with Basal Ganglia Hemorrhage.

| **Table. 1 Demographic Data for Patients with Basal Ganglia Hemorrhage** |  |  |  |  |  |  |
| --- | --- | --- | --- | --- | --- | --- |
| **Variables** | **Total**<br>**(N=387)** | **Surgery**<br>**(n=131)** | **Medical**<br>**(n=255)** | **OR** | **95%CI** | **p-value** |
| **Age Mean±SD** | **58.04±13.01** | 56.86±12.26 | 58.65±13.36 | 0.99 | 0.97-1.00 | 0.200 |
| **Male, n (%)** | **274(70.80)** | 91(33.21) | 183(66.79) | 0.87 | 0.55-1.38 | 0.562 |
| **Female, n (%)** | **113(29.20)** | 41(36.28) | 72(64.72) |  |  |  |
| **CAD, n (%)** | **17(4.39)** | 4(3.03) | 13(5.10) | 0.58 | 0.19-1.82 | 0.352 |
| **CKD, n (%)** | **23(5.94)** | 4(3.03) | 19(7.45) | 0.39 | 0.13-1.17 | 0.092 |
| **Smoking, n (%)** | **130(33.59)** | 44(33.33) | 86(33.73) | 0.98 | 0.52-1.51 | 0.654 |
| **Alcoholic, n (%)** | **175(45.22)** | 61(46.21) | 114(44.71) | 1.06 | 0.70-1.62 | 0.778 |
| **Volume of**<br>**hematoma**<br>**Median(IQR)** | **34.2(14.35,69.85)** | 58(36.2,80) | 22(9,55) | 1.01 | 1.01-1.02 | 0.000* |

SD, standard deviation; IQR, Interquartile range
OR, odds ratio
\* P-value < 0.05

In patients receiving medical treatment alone, mean age of deceased patients was 62.96±15.42, whereas it was 56.92±12.05 for survivors (aOR 0.98; 95%CI 0.94-1.02; p = 0.281). In terms of gender, 183 males (71.76%) underwent only medical treatment, with 46 males (25.14%) dying, compared to 72 females (28.24%), with 27 females (37.50%) dying (aOR 1.68; 95%CI 0.57-4.98; p = 0.349). Regarding personal history, including smoking habits, 86 patients (33.73%), with 23 (31.51%) dying, and 63 (34.62%) surviving (aOR 0.79; 95%CI 0.30-2.07; p = 0.628). Alcohol consumption was observed in 114 patients (44.71%), with 30 (41.10%) deaths, and 84 (46.15%) survivors (aOR 0.97; 95%CI 0.38-2.48; p = 0.942). No significant differences were observed in age, gender, smoking, and alcohol habits. However, underlying diseases such as CAD (11 cases, 15.07% vs. 2 cases, 1.1%; aOR 0.02; 95%CI 0.00-0.10; p = 0.000) and CKD (11 cases (15.07%) vs. 8 cases (4.4%); aOR 0.04-0.70; p = 0.000) were found to have statistically significant differences between deceased and surviving patients. Analysis of hematoma volume revealed a median volume of 79.8 ml (IQR 55,117.25) in deceased patients, contrasting with 16.1 ml (IQR 6.4,27.4) in surviving patients, showing a statistically significant difference (aOR 0.94; 95%CI 0.93-0.96; p=0.000) (Table 2). This suggests that hematoma volume may play a crucial role in predicting patient outcomes in basal ganglia hemorrhage cases treated medically.

**Table 2.** Demographic Data for Patients Undergoing Medical Treatment with Basal Ganglia Hemorrhage.

|  |  |  |  |  |  |  |  |
| --- | --- | --- | --- | --- | --- | --- | --- |
| **Variables** | **Total**<br>**(N=255)** | **Died**<br>**(n=73)** | **Survived**<br>**(n=182)** | **uOR** | **aOR** | **95%CI** | **p-value** |
| **Age Mean±SD** | **58.65±13.36** | 62.96±15.42 | 56.92±12.05 | 0.97 | 0.98 | 0.94-1.02 | 0.281 |
| **Male, n (%)** | **183(71.76)** | 46(25.14) | 137(74.86) | 1.79 | 1.68 | 0.57-4.98 | 0.349 |
| **Female, n (%)** | **72(28.24)** | 27(37.50) | 45(62.50) |  |  |  |  |
| **CAD, n (%)** | **13(5.10)** | 11(15.07) | 2(1.1) | 0.06 | 0.02 | 0.00-0.10 | 0.000* |
| **CKD, n (%)** | **19(7.45)** | 11(15.07) | 8(4.4) | 0.26 | 0.17 | 0.04-0.70 | 0.014* |
| **Smoking, n (%)** | **86(33.73)** | 23(31.51) | 63(34.62) | 1.15 | 0.79 | 0.30-2.07 | 0.628 |
| **Alcoholic, n (%)** | **114(44.71)** | 30(41.1) | 84(46.15) | 1.23 | 0.97 | 0.38-2.48 | 0.942 |
| **Volume of**<br>**hematoma**<br>**Median(IQR)** | **21.2(9,55)** | 79.8(55,117.25) | 16.1(6.4,27.4) | 0.95 | 0.94 | 0.93-0.96 | 0.000* |

SD, standard deviation; IQR, Interquartile range
uOR, unadjusted odds ratio; aOR, adjusted add ratio
\* P-value < 0.05

Focusing on the difference in hematoma volume among patients undergoing medical treatment only, the analysis revealed distinct outcomes based on various volume ranges. In the 0-9.9 ml category, 4 patients (5.97%) died, while 63 (94.03%) survived. For the 10-29.9 ml group, 8 patients (8.99%) died, and 81 (91.01%) survived; however, this difference was not statistically significant when compared with the 0-9.9 ml group (aOR 3.71; 95%CI 0.67-20.59; p=0.133). In the 30-39.9 ml group, 1 patient (5.56%) died and 17 (94.44%) survived, with no statistical significance (aOR 4.23; 95%CI 0.31-58.04; p=0.280). In the 40-49.9 ml group, 4 patients (33.33%) died and 8 (66.67%) survived, indicating statistical significant for mortality (aOR 27.00; 95%CI 3.21-227.45; p = 0.002). The 50-59.9 ml group had 6 patients (60%) died and 4 (40%) survived. (aOR 75.30; 95%CI 8.96-632.48; p=0.000). The 60-69.9 ml group had 8 patients (80%) died and 2 (20%) survived (aOR 256.03; 95%CI 26.17-2505.25; p=0.000), the 70-79.9 ml group had 7 patients (77.78%) died and 2 (22.22%) survived (aOR 251.78; 95%CI 25.41-2494.50; p=0.000); and the group with 80 ml or more had 35 patients (87.5%) died and 5 (12.5%) survived. (aOR 395.13; 95%CI 58.43-2671.83; p=0.000) (Table. 3).

**Table 3.** Analysis of Varying Hematoma Volumes in Patients Undergoing Medical Treatment for Basal Ganglia Hemorrhage.

|  |  |  |  |  |  |  |
| --- | --- | --- | --- | --- | --- | --- |
| **Imaging** |  |  |  | **Died** |  |  |
| **Variables** | **Total**<br>**(N =255)** | **Died**<br>**(n=73)** | **Survived**<br>**(n=182)** | **aOR\*\*** | **95%CI** | **p-value** |
| **Volume(ml)**<br>**, n (%)** |  |  |  |  |  |  |
| **0-9.9** | **67(26.27)** | 4(5.97) | 63(94.03) | - | - | - |
| **10-29.9** | **89(34.90)** | 8(8.99) | 81(91.01) | 3.71 | 0.67-20.59 | 0.133 |
| **30-39.9** | **18(7.06)** | 1(5.56) | 17(94.44) | 4.23 | 0.31-58.04 | 0.280 |
| **40-49.9** | **12(4.71)** | 4(33.33) | 8(66.67) | 27.00 | 3.21-227.45 | 0.002* |
| **50-59.9** | **10(3.92)** | 6(60.00) | 4(40.00) | 75.30 | 8.96-632.48 | 0.000* |
| **60-69.9** | **10(3.92)** | 8(80.00) | 2(20.00) | 256.03 | 26.17-2505.25 | 0.000* |
| **70-79.9** | **9(3.53)** | 7(77.78) | 2(22.22) | 251.78 | 25.41-2494.50 | 0.000* |
| **>=80** | **40(15.69)** | 35(87.5) | 5(12.5) | 395.13 | 58.43-2671.83 | 0.000* |

aOR, adjusted add ratio
\* P-value < 0.05
\*\* adjusted with CAD,CKD

The comparison of survival rates between surgical management and medical treatment revealed significant findings across different hematoma volume groups. In the 0-29.9 ml category, 16 out of 21 individuals (76.19%) who underwent surgical management were alive compared to 142 out of 154 (92.21%) who received medical treatment, demonstrating statistical significance (aOR 0.13; 95%CI 0.03-0.50; p=0.003). For the 30-39.9 ml group, 13 out of 16 individuals (81.25%) in the surgical management group were alive compared to 17 out of 18 (94.44%) in the medical treatment group, although the difference was not statistically significant (aOR 0.24; 95%CI 0.02-2.54; p=0.234). Similarly, in the 40-49.9 ml group, 16 out of 19 individuals (84.21%) in the surgical management group were alive compared to 8 out of 12 (66.67%) in the medical treatment group, with no statistical significance (aOR 2.81; 95%CI 0.39-20.46; p=0.307). The 50-59.9 ml group showed 9 out of 12 individuals (75%) alive in the surgical management group compared to 4 out of 10 (40%) in the medical treatment group, but this difference was not statistically significant (aOR 4.50; 95%CI 0.57-35.52; p=0.154). In the 60-69.9 ml group, 14 out of 16 individuals (87.5%) survived in the surgical management group compared to 2 out of 10 (20%) in the medical treatment group, showing statistical significance (aOR 24.50; 95%CI 2.83-212.40; p=0.004). For the 70-79.9 ml group, 10 out of 14 individuals (71.43%) were alive in the surgical management group compared to 2 out of 9 (22.22%) in the medical treatment group, with a statistical significance (aOR 8.75; 95%CI 1.24-61.68; p=0.029). In the group with 80 ml or more, 21 out of 34 individuals (61.76%) were alive in the surgical management group compared to 7 out of 42 (16.67%) in the medical treatment group, demonstrating statistical significance (aOR 6.81; 95%CI 2.31-20.13; p=0.001) (Table. 4). These results underscore the impact of hematoma volume on survival rates, with surgical management showing varied outcomes across different volume categories.

**Table 4.** Comparison of the Outcomes of Surgical Intervention Versus Medical Treatment in Individuals with Diverse Volumes of Basal Ganglia Hemorrhage.

|  |  |  |  |  |  |  |  |  |
| --- | --- | --- | --- | --- | --- | --- | --- | --- |
| **Imaging** |  | **Surgery (n=132)** |  | **Medical (n=255)** |  | **Survived** |  |  |
| **Variables** | **Total (N=387)** | **Survived (n=99)** | **Died (n=33)** | **Survived (n=182)** | **Died (n=73)** | **aOR** | **95%CI** | **p-value** |
| **Volume(ml)**<br>**, n (%)** |  |  |  |  |  |  |  |  |
| **0-29.9** | **175(45.22)** | 16(76.19) | 5(23.81) | 142(92.21) | 12(7.79) | 0.13 | 0.03-0.50 | 0.003* |
| **30-39.9** | **34(8.79)** | 13(81.25) | 3(18.75) | 17(94.44) | 1(5.56) | 0.24 | 0.02-2.54 | 0.234 |
| **40-49.9** | **31(8.01)** | 16(84.21) | 3(15.79) | 8(66.67) | 4(33.33) | 2.81 | 0.39-20.46 | 0.307 |
| **50-59.9** | **22(5.68)** | 9(75) | 3(25) | 4(40) | 6(60) | 4.50 | 0.57-35.52 | 0.154 |
| **60-69.9** | **26(6.72)** | 14(87.5) | 2(12.5) | 2(20) | 8(80) | 24.50 | 2.83-212.40 | 0.004* |
| **70-79.9** | **23(5.94)** | 10(71.43) | 4(28.57) | 2(22.22) | 7(77.78) | 8.75 | 1.24-61.68 | 0.029* |
| **>=80** | **76(19.64)** | 21(61.76) | 13(38.24) | 7(16.67) | 35(83.33) | 6.81 | 2.31-20.13 | 0.001* |

aOR, adjusted add ratio
\*P-value < 0.05
\*\* adjusted with CAD,CKD

The analysis of post-treatment outcomes for both medical and surgical interventions revealed that among surviving basal ganglia patients, the median length of hospital stay (LOS) was 9 days (IQR 4,20) in total, comprising 16 days (IQR 8,25) for the surgical group and 7 days (IQR 3,15) for the medical group. However, there were no significant differences in this regard in multivariate analysis (aOR 1.01; 95% CI 0.99-1.02; P= 0.514). Regarding post-treatment complications, pneumonia occurred in a total of 71 patients (25.18%), with 44 patients (44.44%) in the surgical group and 27 patients (14.75%) in the medical group alone, showing a statistically significant difference (aOR 2.23; 95% CI 1.04-4.76; P= 0.039). Urinary tract infections (UTI) were observed in 22 patients (7.83%) overall, with 6 (6.12%) in the surgical group and 16 (8.74%) in the medical group alone, although this difference was not statistically significant (aOR 0.59; 95% CI 0.17-2.03; P= 0.403). Septicemia occurred in 8 patients (2.84%) overall, with 5 (5.05%) in the surgical group and 3 (1.64%) in the medical group alone, without statistical significance (aOR 2.06; 95% CI 0.32-13.43; P= 0.446). Cerebral infarction was solely observed in the medical group, affecting 2 patients (1.09%). Following surgical intervention, complications such as rebleeding at the surgical site were noted in 3 patients (3.03%), and post-operative brain edema was observed in 4 patients (4.04%). The Modified Rankin scale (mRS) before treatment revealed a median score of 5 (IQR 5,5) in all surviving patients with basal ganglia hemorrhage. In the surgical group, the median score was 5 (IQR 5,5), whereas in the medical-only group, it was 5 (IQR 3,5), with statistical significance in the analysis (aOR 1.79; 95% CI 1.01-3.16; P=0.045). After treatment, the overall mRS score was 5 (IQR 3,5). In the surgical group, the score remained stable at 5 (IQR 5,5), while in the medical group, there was an improvement in the score to 4 (IQR 3,5), which was statistically significant in multivariate analysis (aOR 1.65; 95% CI 1.12-2.45; P=0.012). The analysis of changes in mRS scores before and after treatment revealed that while the surgical group barely changed with a score of 0 (IQR 0,0), the medical group displayed slight improvement with a score of 0 (IQR 0,1). However, this difference did not achieve statistical significance (aOR 0.75; 95% CI 0.46-1.22; P = 0.246).(Table.5).

**Table 5.**
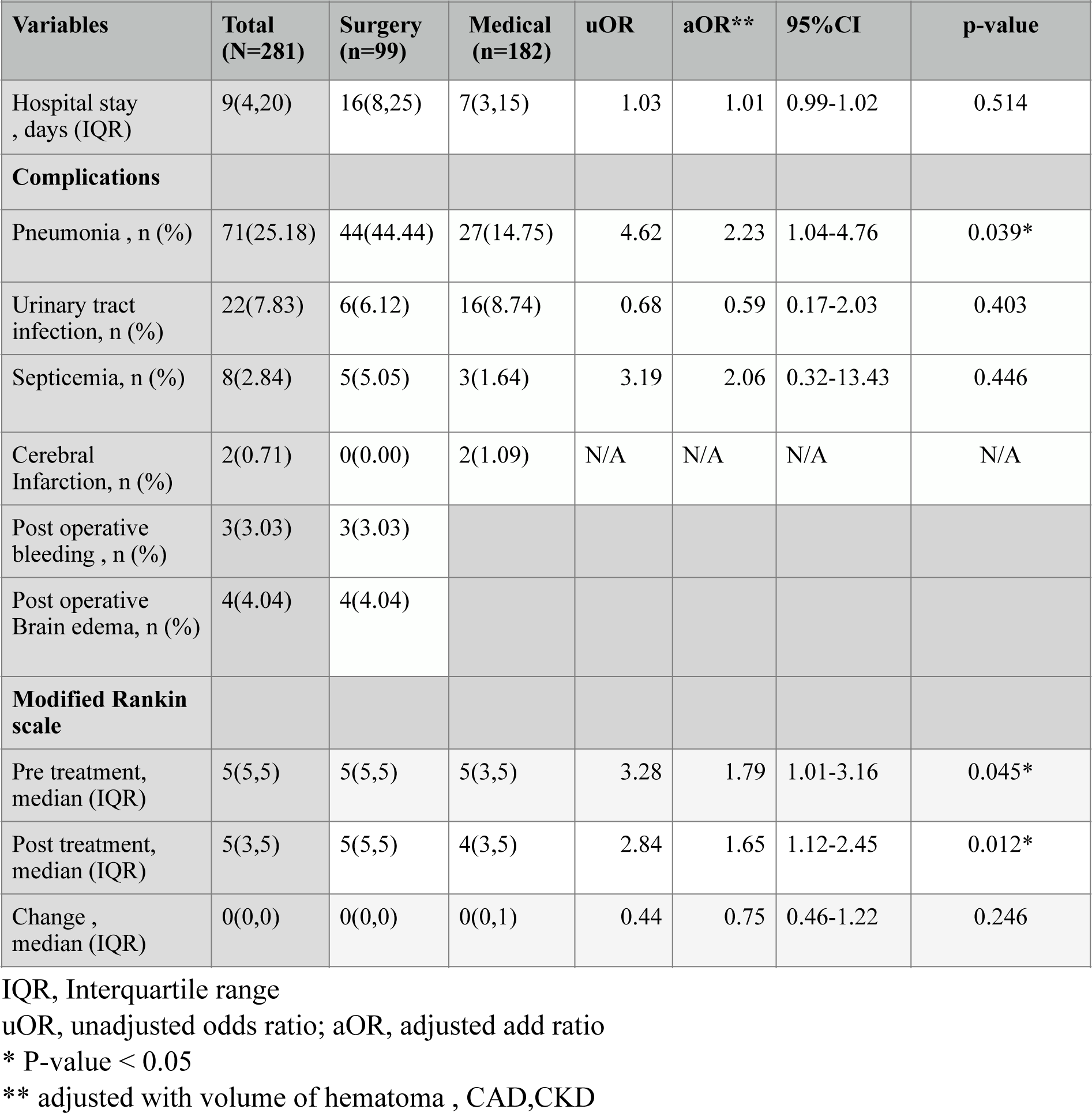
Comparison of Post-Treatment Outcomes Between Surgical Intervention and Medical Treatment in Survived Basal Ganglia Hemorrhage Patients.

|  |  |  |  |  |  |  |  |
| --- | --- | --- | --- | --- | --- | --- | --- |
| **Variables** | **Total**<br>**(N=281)** | **Surgery**<br>**(n=99)** | **Medical**<br>**(n=182)** | **uOR** | **aOR\*\*** | **95%CI** | **p-value** |
| Hospital stay , days (IQR) | 9(4,20) | 16(8,25) | 7(3,15) | 1.03 | 1.01 | 0.99-1.02 | 0.514 |
| **Complications** |  |  |  |  |  |  |  |
| Pneumonia , n (%) | 71(25.18) | 44(44.44) | 27(14.75) | 4.62 | 2.23 | 1.04-4.76 | 0.039* |
| Urinary tract infection, n (%) | 22(7.83) | 6(6.12) | 16(8.74) | 0.68 | 0.59 | 0.17-2.03 | 0.403 |
| Septicemia, n (%) | 8(2.84) | 5(5.05) | 3(1.64) | 3.19 | 2.06 | 0.32-13.43 | 0.446 |
| Cerebral Infarction, n (%) | 2(0.71) | 0(0.00) | 2(1.09) | N/A | N/A | N/A | N/A |
| Post operative bleeding , n (%) | 3(3.03) | 3(3.03) |  |  |  |  |  |
| Post operative Brain edema, n (%) | 4(4.04) | 4(4.04) |  |  |  |  |  |
| **Modified Rankin scale** |  |  |  |  |  |  |  |
| Pre treatment, median (IQR) | 5(5,5) | 5(5,5) | 5(3,5) | 3.28 | 1.79 | 1.01-3.16 | 0.045* |
| Post treatment, median (IQR) | 5(3,5) | 5(5,5) | 4(3,5) | 2.84 | 1.65 | 1.12-2.45 | 0.012* |
| Change , median (IQR) | 0(0,0) | 0(0,0) | 0(0,1) | 0.44 | 0.75 | 0.46-1.22 | 0.246 |

IQR, Interquartile range
uOR, unadjusted odds ratio; aOR, adjusted add ratio
\* P-value < 0.05
\*\* adjusted with volume of hematoma , CAD,CKD

## Discussion

The medical approach to SICH centers around prompt stabilization and addressing the acute condition of intracranial hemorrhage by identifying and treating the root causes of decreased alertness. This encompasses actively rectifying elevated blood pressure and reversing any coagulopathy ^(13)(14)^. In situations where patients display significant or advancing elevation in intracranial pressure (ICP), or if milder symptoms persist despite initial interventions, osmotic therapy is utilized as a therapeutic measure^(15)^. In this study, there were no demographic differences observed between those undergoing surgical intervention and those receiving medical treatment. The multivariate analysis focused on patients exclusively receiving medical treatment, revealing that CKD (aOR 0.17; 95%CI 0.04-0.70), CAD (aOR 0.02; 95%CI 0.00-0.10), and CT brain hematoma volume (aOR 0.94; 95%CI 0.93-0.96) maintained a significant association with an elevated risk of death in cases of basal ganglia hemorrhage (Table 2).

The primary focus of most studies is on patients in a comatose state (with a GCS score <8), those with hematomas exceeding 30 mL, or individuals whose intracranial pressure did not normalize with medical management ^(22, 23, 24)^. While a hematoma volume of at least 30 ml has been considered an indication for surgery, in real-world situations, many patients exhibit tolerance and maintain autoregulation, enabling their survival. The preceding study identified the factors contributing to 30-day in-hospital mortality. It did not include the volume of hematoma but highlighted the association with the degree of compression of the basal cistern and physical examination ^(25)^. Nevertheless, there is no specific volume that can be exclusively managed through medical treatment. In the medical treatment alone group across different hematoma volumes in this study revealed that the 10-39.9 ml group had a non-significantly different likelihood of mortality compared to the less than 10 ml group.

That significant can be concluded that volume 30-39.9 ml group was not absolutely indication for surgical intervention (aOR 4.23; 95%CI 0.31-58.04; p = 0.280) and some patients can be tolerated with ICP autoregulation. However, These findings suggest a clear association between increasing hematoma volume, particularly volumes exceeding 40 ml, and higher mortality rates among patients receiving medical treatment compared to the less than 10 ml group.

The summary suggests that for patients with hemorrhage volumes ranging from 0 to 39.9 ml, treatment solely with medication remains a viable option, with a higher likelihood of survival compared to undergoing surgery. This contradicts previous research findings, which focused on patients with lobar bleedings with volumes greater than 30 ml^(7)^.

Hematoma volume was a crucial factor considered in constructing the Receiver Operating Characteristics (ROC) curve. The primary objective of the ROC curve was to distinguish between patients who survived and those who succumbed to basal ganglia hemorrhage despite undergoing medical treatment. The area under the curve was 0.899 (95% CI 0.85-0.95), and a cutoff value of 45.3 ml was established. This cutoff value effectively predicted survival with medical treatment alone, achieving a sensitivity of 80.82% (95%CI 69.9-89.1), specificity of 91.67% (95%CI 86.8-95.3), Positive likelihood ratio 9.81 (95%CI 5.96-16.13), Negative likelihood ratio 0.21(95%CI 0.13-0.34), Positive predictive value(PPV) 79.7% (95%CI 68.8-88.2) and Negative predictive value(NPV) 92.3% (95%CI 87.4-95.7) (Figure.2). This implies that patients with a hematoma volume of 45.3 ml or less have an 80.82% sensitivity and 91.67% specificity for surviving with only medical treatment.

**Figure.2.**
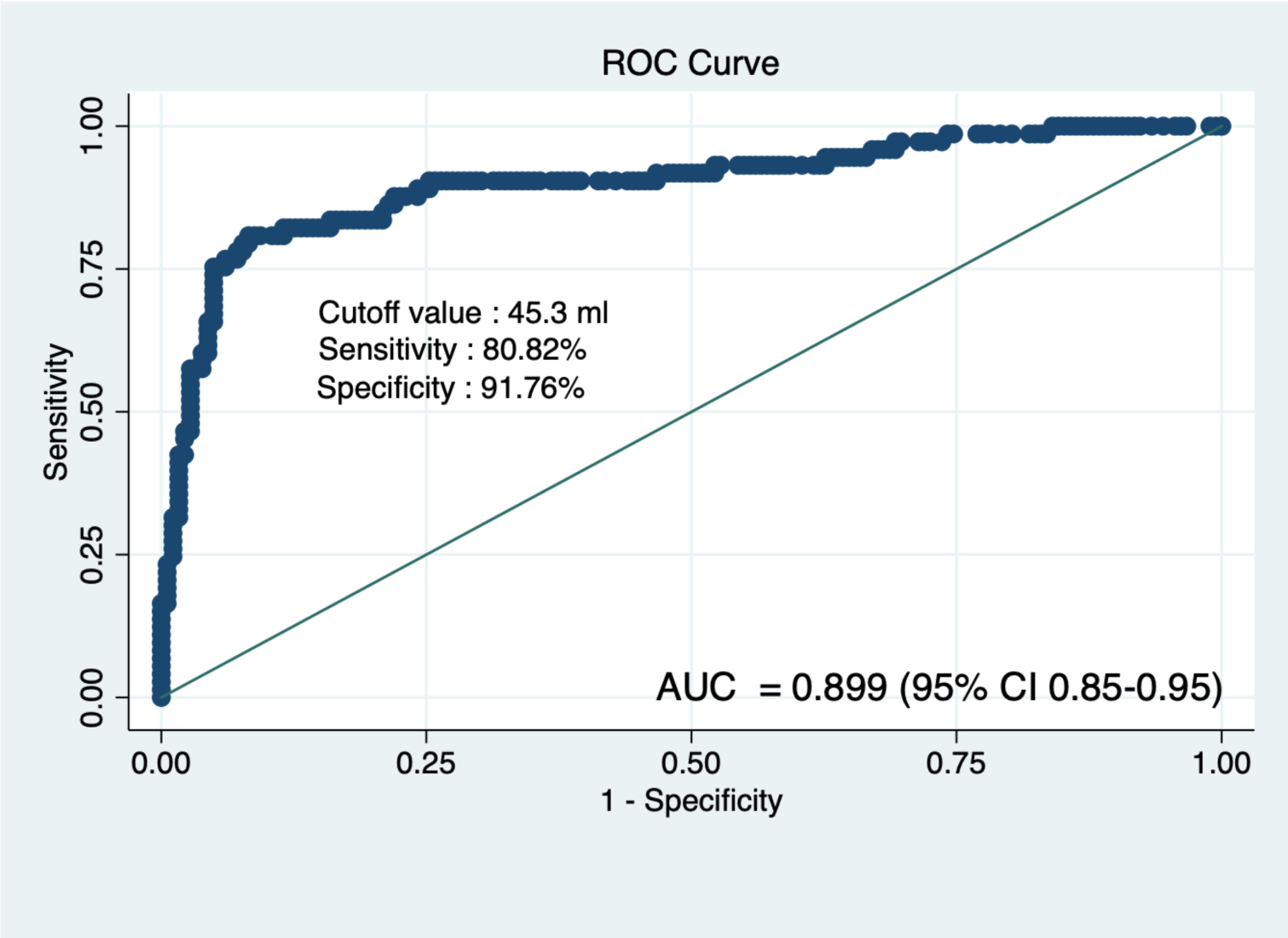
Receiver Operating Characteristic (ROC) curve of hematoma volume in patients undergoing only medical treatment for predicting mortality. AUC mean area under curve; 95% CI, 95% confidence interval

The previous meta-analysis suggests that there is no significant increase in the likelihood of death and dependency at 6 months for patients who undergo surgical treatment (OR 1.23; 95%CI 0.77-1.98) ^(16)^. McKissock et al. indicate a tendency toward a greater risk of death and dependency following surgery (OR 1.20; 95%CI 0.83-1.74)^(16)^. Conversely, an alternative meta-analysis, which compiled data from the studies of Juvela et al. ^(17)^, Auer et al. ^(18)^, and Batjer et al. ^(19)^, demonstrates a trend indicating improved mortality (OR 0.50; 95%CI 0.28-0.92) ^(20)^ associated with surgical intervention. Surgery performed within 8 hours from onset appears to be beneficial^(21)^.. Retrospective case series provide evidence supporting the safety and feasibility of decompressive craniectomy. In this study, it was observed that among patients undergoing surgical intervention with varying hematoma volumes, the surgical group with less than 30 ml exhibited significantly higher mortality than survival (aOR 0.13; 95% CI 0.03-0.50). Additionally, the 30-39.9 ml surgical group showed a trend towards higher mortality than survival, although the difference was not statistically significant (aOR 0.24; 95% CI 0.02-2.54). This suggests that surgical intervention in the group with volumes less than 40 ml may be associated with a significantly higher mortality rate. For the 40-49.9 ml group (aOR 2.81; 95% CI 0.39-20.46) and the 50-59.9 ml surgical group (aOR 4.50; 95% CI 0.57-35.52), there was a tendency suggesting a benefit for surgery in terms of survival over mortality, but these trends were not statistically significant. Despite variations in individual cerebral cortex volume and cranial cavity space, there may be additional factors influencing intracranial pressure within this volume range. Due to the limited population within this subset, further investigation could be warranted. In the surgical group with hematoma volumes of 60 ml or more, there was a notable trend toward increased survival, which was statistically significant. Hematoma volumes of 60 ml or more could potentially increase ICP, which cannot be managed through autoregulation. For a cutoff of 60 ml, the indication for surgery improved the survival rate with a sensitivity of 80.36% (95% CI 67.6-89.8), a specificity of 72.46% (95% CI 60.4-82.5), a positive likelihood ratio of 2.92 (95% CI 1.95-4.37), a negative likelihood ratio of 0.27 (95% CI 0.16-0.47), a PPV of 70.3% (95% CI 57.6-81.1), and a NPV of 82.0% (95% CI 70.0-90.6). Notably, surgical intervention for hematoma volumes surpassing 60 ml also demonstrated a statistically significant benefit in saving lives in cases of basal ganglia hemorrhage.

In a large cohort of patients with ICH, multivariate analysis indicated that a LOS did not significantly correlate with discharge mRS after adjusting for the relevant covariates. The analysis found that worse discharge mRS were significantly associated with older age, greater initial hematoma volume, lower initial GCS, and the need for mechanical ventilation.^(34)^ The paper describes the mean LOS, which was found to be 9.72 days in 76 non-surgery patients and 24 days in 34 surgery patients.^(35)^ Our study reports a non-significant improvement in the length of hospital stay among patients receiving surgical and medical treatment (aOR 1.01; 95%CI 0.99-1.02) (Table.5). Relevant recent papers support and conclude that surgical management does not improve the LOS.

Complications from intracerebral hemorrhage reveal several routes of infection, with pneumonia being the most common disease associated with dependent patients, particularly those with low GCS and high mRS.^(36)^ The increasing odds ratio for pneumonia in the surgery group is significant. This can be explained by several factors, including low scores on independent mRS assessments and a higher prevalence of mechanical ventilation support cases in the surgery group compared to the medical group, as outlined in our paper. Santosh B. Murthy et al. noted a gradual increase in infection rates from 18.7% in 2002–2003 to 24.1% in 2010–2011, with pneumonia being the predominant nosocomial infection at 15.4%, followed by UTI at 7.9%. Our study revealed UTI as the second most prevalent infection, occurring in 7.83% of cases, with no significant disparity between the surgery and medical groups (aOR 0.59; 95% CI 0.17-2.03) (Table.5). Additionally, patients with ICH exhibit increased susceptibility to sepsis due to immunosuppression and dysbiosis of the intestinal microbiota.^(38)^ Sepsis occurs in both the surgery and medical groups with barely any difference, and it is not statistically significant (aOR 2.06; 95% CI 0.32-13.43) (Table.5).

Ruijun Ji et al. developed predictive factors for poor functional outcomes linked to mRS scores ranging from 3 to 6, observed one year after ICH within the derivation cohort. These predictive factors encompass age, admission NIHSS score, GCS, blood glucose levels, ICH location, hematoma volume, and intraventricular hemorrhage^(39)^. The study revealed that both pre-treatment and post-treatment mRS scores in the surgical group were significantly higher than those in the medical-only treatment groups, despite the surgical group exhibiting high hematoma volume and low GCS scores. During the 30-day post-treatment follow-up, a majority of individuals in the surgical group maintained clinical stability (with a median change in mRS of 0 (IQR 0,0)), whereas those undergoing medical treatment showed a slight improvement in mRS (with a median change in mRS of 0(IQR 0,1)) (Table.5). Nevertheless, no statistically significant difference in mRS scores between the two groups was observed.

For the conclusion on morbidity outcomes, it was found that both surgical and medical treatments did not result in significant improvements in mRS scores.

Several papers were developed outlining the criteria for surgery in ICH^(7,29,30,31,32,33)^. Key factors considered included the hematoma volume, GCS, and midline shift. Our observations revealed variations in how the GSC was interpreted by different individuals. The hematoma volume proved advantageous, facilitating earlier decision-making. Consequently, our study highlights hematoma volume as the primary indicator for surgery and conservative treatment, aligning with Abino Luzzi et al.’s systematic reviews, which specify that the surgical volume threshold is at least 60 ml.(Table.6)

**Table 6.** Indications for Surgery as Reported in Various Journals.

| **Table 6. Indications for Surgery as Reported in Various Journals** |  |  |  |  |  |
| --- | --- | --- | --- | --- | --- |
| **Author** | **Intervention** | **Type review** | **Number** | **Year** | **Indication for surgery** |
| Abino Luzzi et al<sup>(7)</sup> | Craniotomy | Systemic review | 19 articles | 2019 | Supratentorial location >60 ml and GCS score 4–8 |
| Parry-Jones AR et al<sup>(29)</sup> | Craniotomy | Retrospective cohort | 860 | 2019 | Volume $\geq 30$ mL or GCS score < 8 |
| Lin F et al<sup>(30)</sup> | Craniotomy | Prospective cohort | 158 | 2022 | Volume $\geq 20$ mL and GCS drop $\geq 2$ |
| khallaf, M et al<sup>(31)</sup> | Craniotomy | Prospective cohort | 66 | 2019 | Volume >30 mL and Midline shift >5 mm or Hydrocephalus |
| Wang, N. Et al<sup>(32)</sup> | Craniotomy | Retrospective cohort | 54 | 2022 | Midline shift >5 mm or Ventricular compression |
| Jiang, L. Et al<sup>(33)</sup> | Endoscopic | Randomize control trial | 91 | 2024 | Neurodeficit and Volume > 20-30 ml |
| Our study | Craniotomy | Retrospective cohort | 387 | 2024 | Volume $\geq 60$ ml, regardless of the GCS<br>Volume 45.3-59.9 ml, depend on the patient's status<br>Volume < 30 ml, not recommended for surgery |

GCS, Glasgow Coma Scale

Currently, machine learning methods such as artificial intelligence (AI) and deep learning are crucial for detecting and classifying intracranial hemorrhage ^(26, 27, 28)^, facilitating early identification and improving diagnostic precision. In the future, AI technologies have the potential to expand capabilities globally, with this study aiding physicians in comprehending prognosis, post-treatment outcomes and facilitating prompt decision-making regarding initial management at healthcare facilities. This advancement can lead to cost reductions linked to referrals or unnecessary treatments.

There are several limitations to consider. Firstly, there is a limitation regarding the follow-up duration, as it extends beyond 30 days without the mRS in assessing long-term outcomes. Secondly, the population included in the study experienced hematoma volume expansion, which was not addressed in this paper. This expansion was observed in 19 cases, comprising 4.68% of the total basal ganglia patient population. However, it’s important to note that the cutoff point for conservative treatment, with a high specificity of 91.67%, was not affected by this exclusion. Lastly, the statistical analysis may be impacted by the presence of elderly patients who exhibit a higher degree of brain atrophy compared to the general population. This factor could influence survival rates in cases of high-volume hematoma without surgical intervention.

## Conclusions

In conclusion surgical intervention demonstrates efficacy in basal ganglia hemorrhage, particularly for cases involving larger hematoma volumes. The application of only medical treatment proves feasible for hematoma volumes within the 0 to 45.3 ml range, with a sensitivity of 80.82% and specificity of 91.67%. The decision to opt for surgical intervention in cases with volumes between 45.3 ml and 59.9 ml depends on the surgeon’s discretion and various contributing factors. However, hematoma volumes of at least 60 ml clearly indicate the necessity for surgical intervention in patients with basal ganglia hemorrhage. Nonetheless, there is no improvement in mRS within 30 days with either medical and surgical treatments.

## Article information

## Data Availability

The data that support the findings of this study are available from the first author, where data were recorded in Google Drive. The data supporting the findings of this study are available upon reasonable request via email at.

-

## Nonstandard Abbreviations and Acronyms

AI: Artificial intelligence
aOR: Adjusted odd ratios
CKD: Chronic kidney disease
CAD: Coronary heart disease
GSC: Glasgow Coma Scale
ICP: Intracranial pressure
IQR: Interquartile range
LOS: Length of hospital stay
mRS: Modified Rankin scale
NPV: Negative predictive value
PPV: Positive predictive value
ROC: Receiver Operating Characteristics
SICH: Spontaneous intracerebral hemorrhage
uOR: Unadjusted odd ratios

